# Head-to-head comparison of nasal and nasopharyngeal sampling using SARS-CoV-2 rapid antigen testing in Lesotho

**DOI:** 10.1101/2021.12.29.21268505

**Authors:** Niklaus D Labhardt, Lucia González Fernández, Bulemba Katende, Josephine Muhairwe, Moniek Bresser, Alain Amstutz, Tracy R Glass, Morten Ruhwald, Jilian A Sacks, Camille Escadafal, Mathabo Mareka, Mooko Sekhele Mookho, Margaretha de Vos, Klaus Reither

## Abstract

**Objectives:** To assess the real-world diagnostic performance of nasal and nasopharyngeal swabs for SD Biosensor STANDARD Q COVID-19 Antigen Rapid Diagnostic Test (Ag-RDT).

**Methods:** Individuals ≥5 years with COVID-19 compatible symptoms or history of exposure to SARS-CoV-2 presenting at hospitals in Lesotho received two nasopharyngeal and one nasal swab. Ag-RDT from nasal and nasopharyngeal swabs were performed as point-of-care on site, the second nasopharyngeal swab used for polymerase chain reaction (PCR) as the reference standard.

**Results:** Out of 2198 participants enrolled, 2131 had a valid PCR result (61% female, median age 41 years, 8% children), 84.5% were symptomatic. Overall PCR positivity rate was 5.8%. The sensitivity for nasopharyngeal, nasal, and combined nasal and nasopharyngeal Ag-RDT result was 70.2% (95%CI: 61.3-78.0), 67.3% (57.3-76.3) and 74.4% (65.5-82.0), respectively. The respective specificity was 97.9% (97.1-98.4), 97.9% (97.2-98.5) and 97.5% (96.7-98.2). For both sampling modalities, sensitivity was higher in participants with symptom duration ≤ 3days versus ≤ 7days. Agreement between nasal and nasopharyngeal Ag-RDT was 99.4%.

**Conclusions:** The STANDARD Q Ag-RDT showed high specificity. Sensitivity was, however, below the WHO recommended minimum requirement of ≥ 80%. The high agreement between nasal and nasopharyngeal sampling suggests that for Ag-RDT nasal sampling is a good alternative to nasopharyngeal sampling.

**Highlights:**

- Prospective study on real-world diagnostic performance of nasal and nasopharyngeal SD Biosensor STANDARD Q COVID-19 Ag Test in 2131 participants in a rural African setting
- The sensitivity of the STANDARD Q COVID-19 Ag Test was below the World Health Organization requirement of ≥ 80% but met the specificity requirement of ≥97%.
- Sensitivity was higher in the following subpopulations: persons with symptoms ≤3 days, and Ct value < 25.
- In head-to-head comparison nasal and nasopharyngeal sampling had comparable sensitivity and specificity and an overall test agreement of 99.4%, indicating that the more convenient nasal sampling could be used for SARS-CoV-2 rapid antigen tests.
- 24 of the 2131 participants with COVID-19 symptoms had pulmonary tuberculosis with a positive Xpert Ultra test on sputum.

## Introduction

Wide-spread testing and contact tracing remain key to contain coronavirus disease 2019 (COVID-19) outbreaks, particularly in settings with low vaccination coverage(1). In many sub-Saharan African countries, health systems are struggling to meet the demand of Severe Acute Respiratory Syndrome Coronavirus-2 (SARS-CoV-2) testing due to limited technical and human resource capacity to perform nucleic acid amplification tests, such as real-time Reverse Transcription-Polymerase Chain Reaction (PCR) testing at larger scale(2). SARS-CoV-2 antigen rapid diagnostic tests (Ag-RDTs) that can be used at point-of-care by health professionals, as well as trained lay workers could replace more resource intensive and technical demanding PCR testing in these settings(3).

The WHO recommends the use of Ag-RDTs that have a sensitivity ≥80% and a specificity ≥97% for testing symptomatic and asymptomatic individuals at high risk of infection, including contacts and health workers, for SARS-CoV-2 infection, particularly in settings where PCR testing capacity is limited(4). A living systematic review and meta-analysis on the accuracy of Ag-RDTs, last updated in August 2021 with 133 clinical studies included, found a pooled Ag-RDT sensitivity and specificity of 71.2% and 98.9%, respectively(5). Test accuracy varied between studies using Ag-RDTs from different manufacturers, different sampling strategies and different cycle thresholds (Ct) used for the reference PCR. Only 2 studies included in this review were conducted in sub-Saharan Africa (6,7). Both used nasopharyngeal sampling, one applied the SD Biosensor STANDARD Q COVID-19 Ag Test, the other the Panbio ™ COVID-19 Ag Rapid Test.

Whereas nasopharyngeal sampling is generally considered safe and may rarely lead to severe complications, it is usually very unpleasant to the patient (8). Alternative sampling methods, including saliva, oropharyngeal swabs, or nasal swabs, either self-collected or collected by trained operators-, have been proposed to increase acceptance of SARS-CoV-2 testing. A review over 23 studies, none from Africa, concluded that for PCR, nasal and saliva sampling were accurate and clinically acceptable alternatives in outpatient settings(9). For rapid antigen tests, comparable sensitivity has been reported for nasopharyngeal and nasal mid-turbinate swabs(10,11).

For remote, resource-limited settings, it is important to identify diagnostically accurate, easily applicable, safe, and convenient SARS-CoV-2 testing strategies that can be provided by non-professional health cadres and that are accepted by the community. We here report diagnostic performance of nasopharyngeal and nasal sampled Ag-RDTs (Biosensor STANDARD Q COVID-19 Ag Test) compared to PCR from nasopharyngeal samples for diagnosis of SARS-CoV-2 in symptomatic and asymptomatic contact persons at rural hospitals in Lesotho, Southern Africa.

## Materials and Methods

### Study design

This study is part of the project *Mitigation Strategies for Communities With COVID-19 Transmission in Lesotho* (MistraL). The project started in October 2020 at two rural district hospitals in Northern Lesotho aiming to support Lesotho’s health system in SARS-CoV-2 screening and diagnosis (https://brc.ch/research/mistral/). To assess diagnostic accuracy of the STANDARD Q COVID-19 Ag Test (SD Biosensor, Republic of Korea) with provider-collected anterior nasal and nasopharyngeal sampling (index tests), compared to PCR from nasopharyngeal sampling (reference test), we prospectively and consecutively enrolled adults and children presenting symptoms compatible with COVID-19 and/or with a history of SARS-CoV-2 exposure.

### Setting and participants

The study was conducted in Northern Lesotho from 28.12.2020 to 30.09.2021 at the Government District Hospital of Mokhotlong and the St Charles Missionary Hospital Seboche and additionally from 21.01.2021 to 12.02.2021 at the Butha-Buthe Government District Hospital. These hospitals serve a population of about 220’000, mainly living in rural communities scattered over a mountainous area in Northern Lesotho. During the 10 months of the study implementation, the Ministry of Health of Lesotho reported the surge of three waves of increased incidence of COVID-19 cases and implemented social mobility restrictions and lockdowns of different intensity. Lesotho’s first COVID-19 wave started in December 2020, the second in May 2021 and the third in September 2021(12).

As part of routine procedures, all children ≥ 5 years, adolescents, and adults attending health services at one of the hospitals were pre-screened for COVID-19 related symptoms at the entrance gate. Any person with body temperature ≥38°C (non-contact forehead thermometer) or reporting at least one out of the following 10 symptoms was eligible for SARS-CoV-2 testing: fever/chills, cough, tiredness, dyspnea, sore throat, body pain, diarrhea, loss of taste/smell, recent weight loss, night sweats. Further eligible were individuals reporting close contact to a probable or confirmed COVID-19 case in the last 14 days, defined as contact <1m for ≥15 min, direct physical contact, or direct care without appropriate personal protective equipment. Informed consent was obtained after pre-screening. A focused medical history was obtained, and a clinical examination was performed on each subject enrolled in the study. Individuals who were deemed critically ill by the healthcare provider, i.e., adults with altered mental status, tachypnea (≥22/min), SpO2<94%, or systolic blood pressure <100mgHg, and children with signs of pneumonia and central cyanosis, SpO2<94%, general danger signs, or tachypnea (5-9 yrs ≥ 30/min, ≥10 yrs ≥ 20/min) were assessed by a hospital physician, who decided whether the participant could remain in the study for all investigations or immediately referred to emergency care.

### Procedures and test methods

For SARS-CoV-2 diagnosis, a study nurse consecutively collected three samples from each participant in the following order: anterior nasal swab for Ag-RDT (Mokhotlong and Seboche site only), nasopharyngeal swab for PCR and nasopharyngeal swab for Ag-RDT. A posterior-anterior chest x-ray was also performed in all adult non-pregnant participants with at least one symptom compatible with COVID-19 (see above), or with clinical signs of tuberculosis or upon specific request by the physician in case of children and pregnant women. Participants with clinical and/or radiological suspicion of tuberculosis were referred to the tuberculosis department for further work-up. Additionally, study participants that were eligible for HIV testing according to Lesotho National Guidelines (unknown HIV status, last negative HIV test ≥12 months ago, high risk for HIV infection, or recent risk for exposure) were offered on-spot HIV testing and referred for to the HIV department in case of a positive test result. Due to logistic reasons, the Butha-Buthe site did not perform nasal swaps but followed all other study procedures as mentioned above.

For SARS-CoV-2 Ag-RDT, we used the STANDARD Q COVID-19 Ag Test commercial test kits (SD Biosensor, Republic of Korea) in both nasopharyngeal and the nasal samples., The Ag-RDTs were performed by nurses directly after sampling following the manufacturer’s instructions (13). STANDARD Q COVID-19 Ag Test is a chromatographic immunoassay targeting the SARS-CoV-2 nucleocapsid antigen. According to the manufacturer, it has a sensitivity and specificity of 84.97% and 98.94% respectively(14). The commercial SD Biosensor STANDARD Q COVID-19 Ag Test kits for nasal and nasopharyngeal only differ by the sterile swab for sample collection which is longer and thinner in the case of the nasopharyngeal kit and shorter and thicker in the case of the nasal kit. Prior to sample collection, participants had to blow their nose. For the nasal sample collection, once the participant was seated, with their head back slightly, the swab was inserted while rotating it in each nostril, gently pushing the swab up to about 2 cm into the nostril until resistance was met and rotated for 3-5 seconds. Nasopharyngeal sample collection was done by inserting the nasopharyngeal swab in horizontal position into the back of the nasopharynx until resistance was felt, while rotating the swab at least 5 times. The test result was read by a trained heath professional after 15-30 minutes using visual inspection (not a digital reader or a digital application). The Ag-RDT test result was reported positive if both the control line and the SARS-Cov-2 Ag test line were present. The test result was reported negative if only the control line was present. The test result was invalid if the control line was missing.

As a reference standard, the first nasopharyngeal swab was sent within 24 hours to the Lesotho National Reference Laboratory, where PCR was performed within a maximum period of 72 hours after sample collection. Before and during shipment, samples were kept at a temperature between 4°C and 8°C. PCRs were performed on the ABI 7500 Real-Time PCR platform (Applied Biosystem, USA) targeting the N (nucleocapsid) Gene and ORF1ab (open reading frame) Gene following an in-house protocol. At certain days, when there was low throughput, the National Reference Laboratory tested the samples using the Xpert Xpress SARS-CoV-2 (Cepheid, USA) assay instead. Xpert Xpress targets the N2 (nucleocapsid) Gene and the E (envelope) Gene. Specialized laboratory technicians performed the PCRs according to the manufacturer’s standard operating procedures. Laboratory technicians were not aware of the participant’s Ag-RDT result. For the samples tested using the ABI 7500 PCR platform, a detectable SARS-CoV-2 below a Ct value of 35 in both targets, N Gene and ORF1ab gene, were categorized as positive. PCRs with both N Gene and ORF1ab Ct value ≥ 35 or no Ct value for any gene were categorized as negative. Discrepant results where Ct value for one target was <35 and ≥35 for the other one, were considered indeterminate. For Xpert Xpress SARS-CoV-2 reporting followed the manufacturer’s instructions where detection of N2 target was categorized positive, detection of E in the absence of N2 as indeterminate and no detection of both targets but valid control as negative. Regardless the PCR platform used, samples with indeterminate and invalid results were repeated once.

### Data collection and analysis

At both sites, trained study personnel collected participants’ information and results from the Ag-RDT into an Open Data Kit (ODK) database. PCR results from the National Reference Laboratory were reported on specific paper-based study forms, and, at a later stage entered into the study database. The data-manager performed weekly checks on data consistency and completeness, the study team in Lesotho manually checked and clarified all queries.

Participants’ clinical and demographic characteristics are reported descriptively, using median with inter-quartile ranges (IQR) and proportions as appropriate. Sensitivities and specificities were calculated in two-by-two tables using the PCR result as reference standard. We conducted three main analyses on three different index tests: nasopharyngeal Ag-RDT alone, nasal Ag-RDT alone and nasopharyngeal and nasal RDT combined and then assessed test agreement between nasal and nasopharyngeal sampling. We further performed subgroup analyses for sensitivity and specificity of nasal and nasopharyngeal Ag-RDT according to different Ct values for the N Gene (<20, 20-24, 25-29, 30-34) as well as according to symptom duration (≤3 days and ≤7 days). For the analysis stratified by Ct-value only samples processed on the ABI platform were included, because a standardized conversion of Ct values from different platforms into viral load, e.g. using serial of dilutions of cultured SARS-CoV-2, was not possible in our setting(15). Only participant records with a valid PCR result, i.e. positive or negative, were included in the analysis.

### Ethics

This study was approved by the Ethic Committee Switzerland (Ethikkommission Nordwest-und Zentralschweiz (EKNZ) AO_2020-00018) and the National Health and Research and Ethics Committee of Lesotho (NH-REC; ID-107-2020). All adult participants provided written informed consent. In case of illiteracy, the adult participant signed with a thumbprint and an independent person signed as a witness. Participants older than 7 and younger than 18 years provided written assent, in addition to a written consent from the caregiver. For participants above 5 and below 7 years of age, the caregiver provided a written consent only.

## Results

Among 2198 included participants, 2131 had a valid PCR result, 26 (1.2%) of PCR results were indeterminate or invalid, and 41 (1.9%) were missing. Regarding Ag-RDTs, 2198 were eligible for nasopharyngeal results (all 3 sites) and 2093 for a nasal Ag-RDT result (Mokhotlong and Seboche site). Nasopharyngeal Ag-RDT were missing for 46/2198 (2.1%) and nasal Ag-RDT were missing for 88/2093 (4.2%) participants. None of the Ag-RDT results were reported invalid, resulting in 2126 participants with nasopharyngeal Ag-RDT and PCR results, 1989 with nasal Ag-RDT and PCR result and 1986 who had valid results from all three tests (Figure 1).

**Figure 1:**
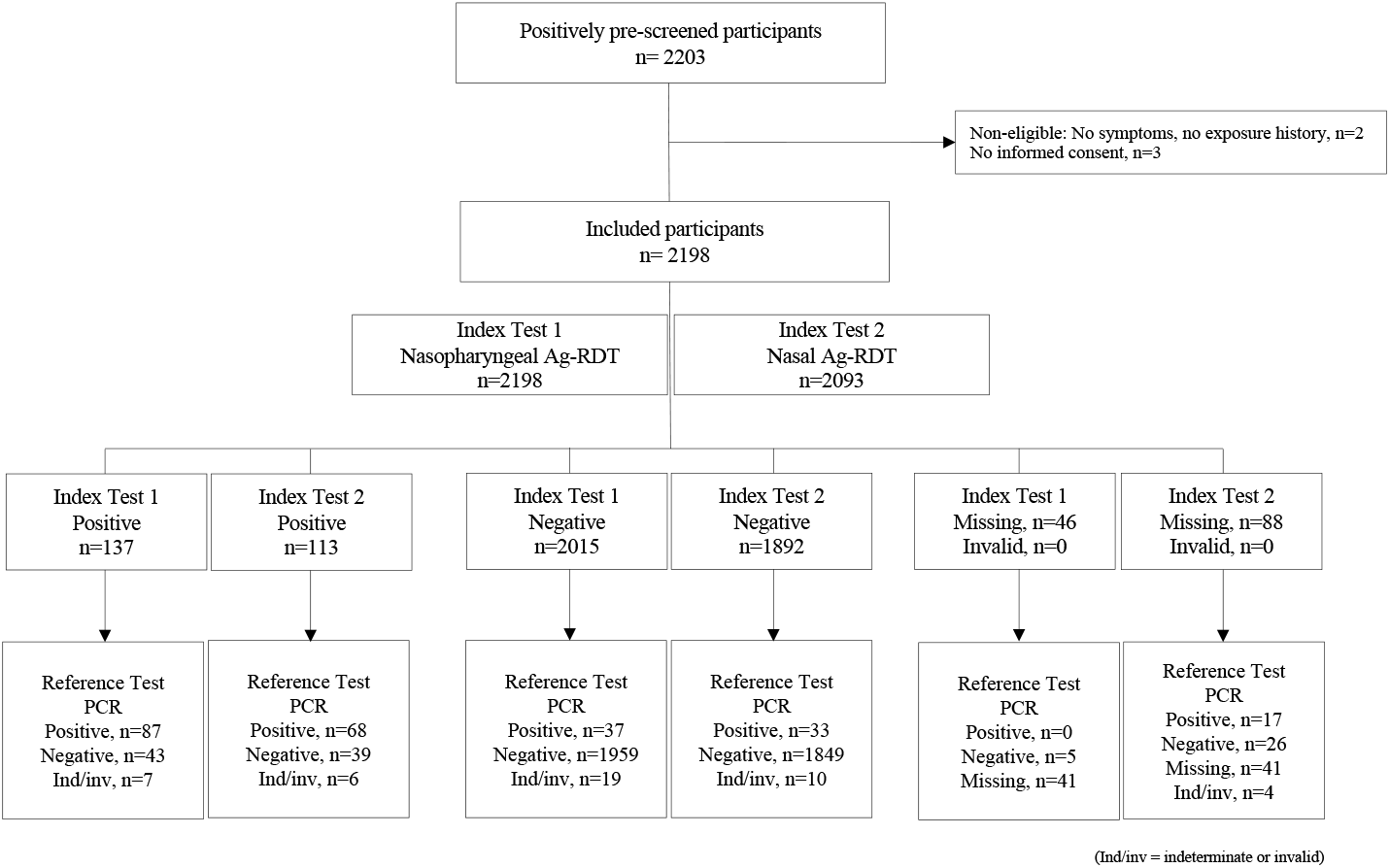
STARD Diagram of the study

Characteristics of the 2131 participants with a valid SARS-CoV-2 PCR result are displayed in table 1. The median age was 41 years (IQR 28-60), 61% (1224) were female, and 8.8% (176) were children or adolescents between 5 to 18 years old. Among the included participants, 1800 (84.5%) reported at least one symptom compatible with COVID-19, 181 (8.5%) reported no symptoms and 150 (7.0%) had missing symptom information. Regarding SARS-CoV-2 contact, 35 (1.6%) reported recent exposure but no symptoms, 43 (2.0%) exposure and symptoms. Overall, 21.2% (452/2131) were HIV positive, 10.3% (191/2131) reported past TB history and 10.9% (232/2131) reported another known chronic comorbidity.

**Table 1:** Demographic and clinical characteristics of study participants

|  | Negative PCR Test | Positive PCR Test | Total |
| --- | --- | --- | --- |
| Total number with a valid PCR result | 2007 (100%) | 124 (100%) | 2131 (100%) |
| Gender |  |  |  |
| Female | 1224 (61.0%) | 70 (56.5%) | 1294 (60.7%) |
| Male | 783 (39.0%) | 54 (43.5%) | 837 (39.3%) |
| Age, years (IQR) | 41 (28-60) | 41 (29-58) | 41 (28-60) |
| Child/adolescent (<18 years) | 176 (8.8%) | 2 (1.6%) | 178 (8.4%) |
| Adult (≥18 years) | 1831 (91.2%) | 122 (98.4%) | 1953 (91.6%) |
| SARS-CoV-2 contact in past 14 days |  |  |  |
| Yes | 68 (4.8%) | 11 (10.8%) | 79 (5.2%) |
| No | 563 (40.1%) | 46 (45.1%) | 609 (40.5%) |
| Unknown | 772 (55.0%) | 45 (44.1%) | 817 (54.3%) |
| **Symptoms** |  |  |  |
| Time since onset of symptoms (Median time, IQR) | 4 (2-7) | 4 (2-6) | 4 (2-7) |
| Fever | 8 (0.5%) | 1 (0.9%) | 9 (0.5%) |
| Cough | 1026 (59.5%) | 84 (75.0%) | 1110 (60.5%) |
| Pronounced tiredness | 483 (29.3%) | 50 (45.0%) | 533 (30.2%) |
| Shortness of breath | 175 (11.0%) | 12 (11.1%) | 187 (11.1%) |
| Chest pain | 138 (7.4%) | 10 (8.6%) | 148 (7.5%) |
| Fatigue | 379 (20.3%) | 46 (39.7%) | 425 (21.5%) |
| Sore throat | 298 (18.5%) | 16 (15.1%) | 314 (18.3%) |
| Muscle pain | 151 (8.1%) | 21 (18.1%) | 172 (8.7%) |
| Diarrhoea | 107 (6.9%) | 7 (6.6%) | 114 (6.8%) |
| Loss of sense or smell | 129 (8.2%) | 19 (17.6%) | 148 (8.8%) |
| Runny nose | 81 (4.3%) | 8 (6.9%) | 89 (4.5%) |
| Headache | 193 (10.3%) | 24 (20.7%) | 217 (11.0%) |
| Skin rash | 8 (0.4%) | 0 (0.0%) | 8 (0.4%) |
| Weight loss | 119 (7.6%) | 7 (6.6%) | 126 (7.5%) |
| Night sweats | 105 (6.7%) | 12 (11.0%) | 117 (7.0%) |
| Vomiting | 42 (2.3%) | 3 (2.6%) | 45 (2.3%) |
| Failure to thrive | 0 (0.0%) | 0 (0.0%) | 0 (0.0%) |
| Poor appetite | 1 (2.9%) | 0 (0.0%) | 1 (2.9%) |
| **History of tuberculosis** |  |  |  |
| Yes | 184 (10.6%) | 7 (6.4%) | 191 (10.3%) |
| No | 1550 (89.1%) | 103 (93.6%) | 1653 (89.4%) |
| Unknown | 6 (0.3%) | 0 (0.0%) | 6 (0.3%) |
| **Miner (adults only)** |  |  |  |
| Yes | 110 (8.2%) | 10 (9.9%) | 120 (8.4%) |
| No | 1220 (91.5%) | 91 (90.1%) | 1311 (91.4%) |
| **Ever smoked (adults only)** |  |  |  |
| Yes | 100 (5.0%) | 10 (8.1%) | 110 (5.2%) |
| Current smoker | 84 (84.0%) | 9 (90.0%) | 93 (84.5%) |
| No | 1231 (92.5%) | 91 (90.1%) | 1322 (92.3%) |
| **Alcohol (adults only)** |  |  |  |
| No | 1156 (86.7%) | 80 (79.2%) | 1236 (86.1%) |
| Yes | 178 (13.3%) | 21 (20.8%) | 199 (13.9%) |
| Drinks per day |  |  |  |
| 1-2 | 57 (32.0%) | 5 (23.8%) | 62 (31.2%) |
| 3-4 | 53 (29.8%) | 2 (9.5%) | 55 (27.6%) |
| 5-6 | 46 (25.8%) | 10 (47.6%) | 56 (28.1%) |
| ≥ 7 | 22 (12.4%) | 4 (19.0%) | 26 (13.1%) |
| Binge drinking (≥6 drinks) |  |  |  |
| Never | 23 (12.9%) | 0 (0.0%) | 23 (11.6%) |
| Monthly or less | 122 (68.5%) | 13 (61.9%) | 135 (67.8%) |
| Weekly or more | 33 (18.5%) | 8 (38.1%) | 41 (20.6%) |
| **HIV status** |  |  |  |
| Positive | 437 (21.7%) | 15 (12.1%) | 452 (21.2%) |
| Taking ART | 409 (93.6%) | 14 (93.3%) | 423 (93.6%) |
| Newly tested | 15 (3.4%) | 0 (0.0%) | 15 (3.3%) |
| Negative | 1083 (54.0%) | 82 (66.1%) | 1165 (54.7%) |
| Refused to answer/test | 8 (0.4%) | 3 (2.4%) | 11 (0.5%) |
| Unknown | 479 (23.9%) | 24 (19.4%) | 503 (23.6%) |
| **Concomitant diseases (reported)** |  |  |  |
| Diabetes | 33 (2.4%) | 7 (6.9%) | 40 (2.7%) |
| Hypertension | 165 (11.8%) | 16 (15.7%) | 181 (12.0%) |
| Heart disease | 7 (0.5%) | 0 (0.0%) | 7 (0.5%) |
| Kidney disease | 0 (0.0%) | 0 (0.0%) | 0 (0.0%) |
| Malnutrition | 0 (0.0%) | 0 (0.0%) | 0 (0.0%) |
| Cancer | 0 (0.0%) | 0 (0.0%) | 0 (0.0%) |
| Chest-lung disease | 11 (0.5%) | 0 (0.0%) | 11 (0.5%) |

(Percentages are of non-missing data)

The overall SARS-CoV-2 positivity rate was 5.8% (124/2131), 6.5% (137/2152) and 5.8% (137/2005) for PCR, nasopharyngeal Ag-RDT and nasal Ag-RDT. The sensitivity for nasopharyngeal, nasal Ag-RDT, and the combination of results from nasal and nasopharyngeal Ag-RDT was 70.2% (95%CI: 61.3-78.0), 67.3% (57.3-76.3) and 74.4% (65.5-82.0), respectively. The respective specificity was 97.9% (97.1-98.4), 97.9% (97.2-98.5) and 97.5% (96.7-98.2), the respective positive predictive value was 66.9%, 63.6% and 64.9%, and the respective negative predictive value was 98.1%, 98.2% and 98.4%.

The overall agreement between nasal and nasopharyngeal Ag-RDT was 99.4%, with 103 tests both positive, 1871 tests both negative, only nasopharyngeal Ag-RDT positive in 8 subjects, and only nasal Ag-RDT positive in 4 subjects.

### Subgroup analyses

The performance of the nasopharyngeal and the nasal Ag-RDTs was in agreement with regards to symptom duration and Ct value for N gene (Figure 2). Figure 3 displays overall sensitivity for the different Ct value threshold categories. Sensitivity was highest for Ct values between 20-24. Accordingly, most false-negative nasal and nasopharyngeal Ag-RDT test results were found in samples from participants with high Ct value (i.e. ≥25).

**Figure 2:**
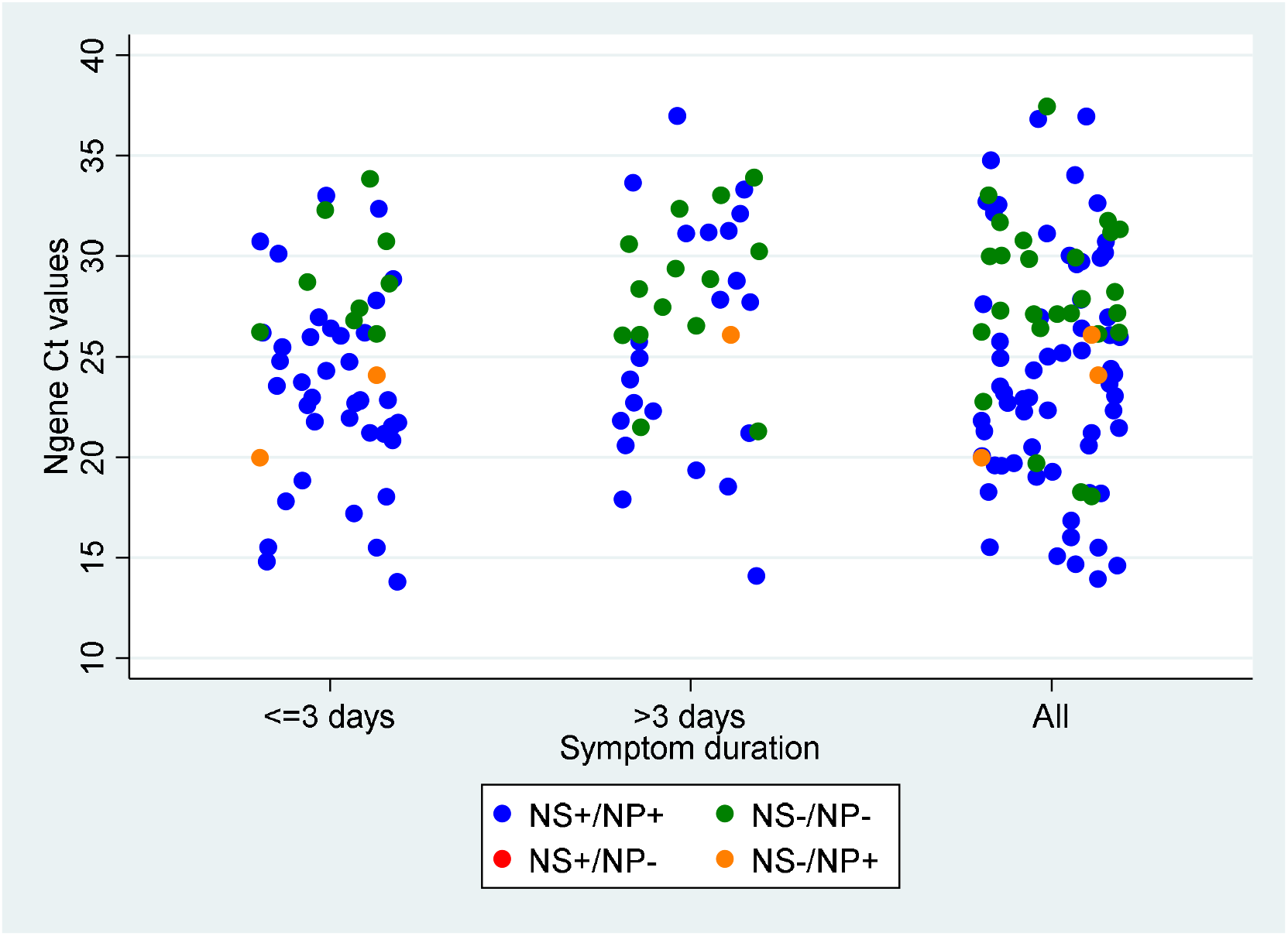
N gene Ct values by the duration of any symptom recorded and according to the agreement of the Ag-RDTs; NS nasal sampling; NP nasopharyngeal sampling

**Figure 3:**
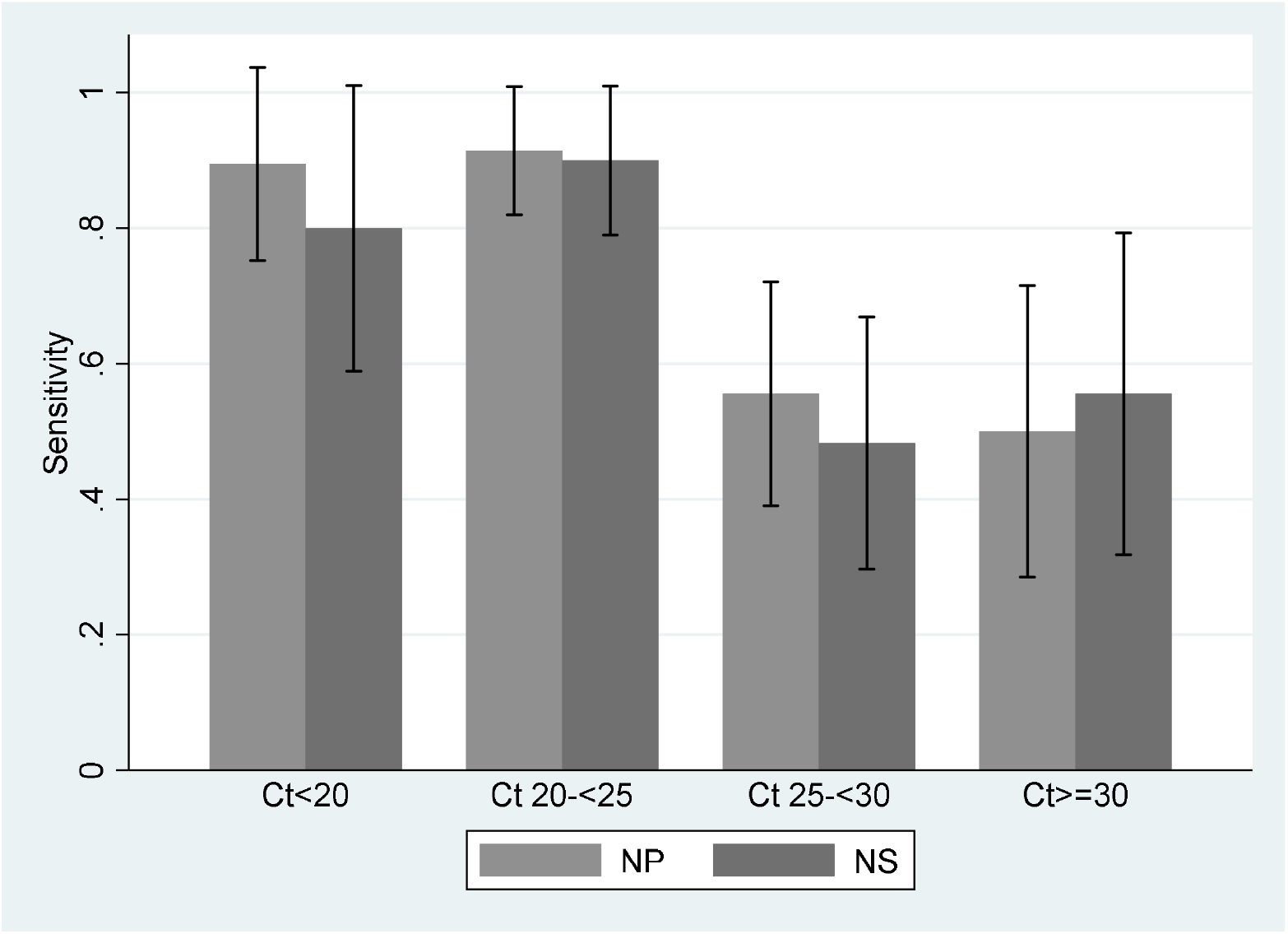
Sensitivity of nasal Ag-RDT (NS) and nasopharyngeal Ag-RDT (NP) according to PCR Ct value.

Sensitivity and specificity stratified by symptoms and symptom duration are displayed in table 2. Sensitivity of both, nasal and nasopharyngeal Ag-RDT was higher in participants with symptom duration ≤ 3 days as compared to ≤ 7 days. Among the 178 children 5 to 18 years, 2 (1%) had a positive PCR test. Due to the low number of cases no subgroup analysis was performed.

**Table 2:**
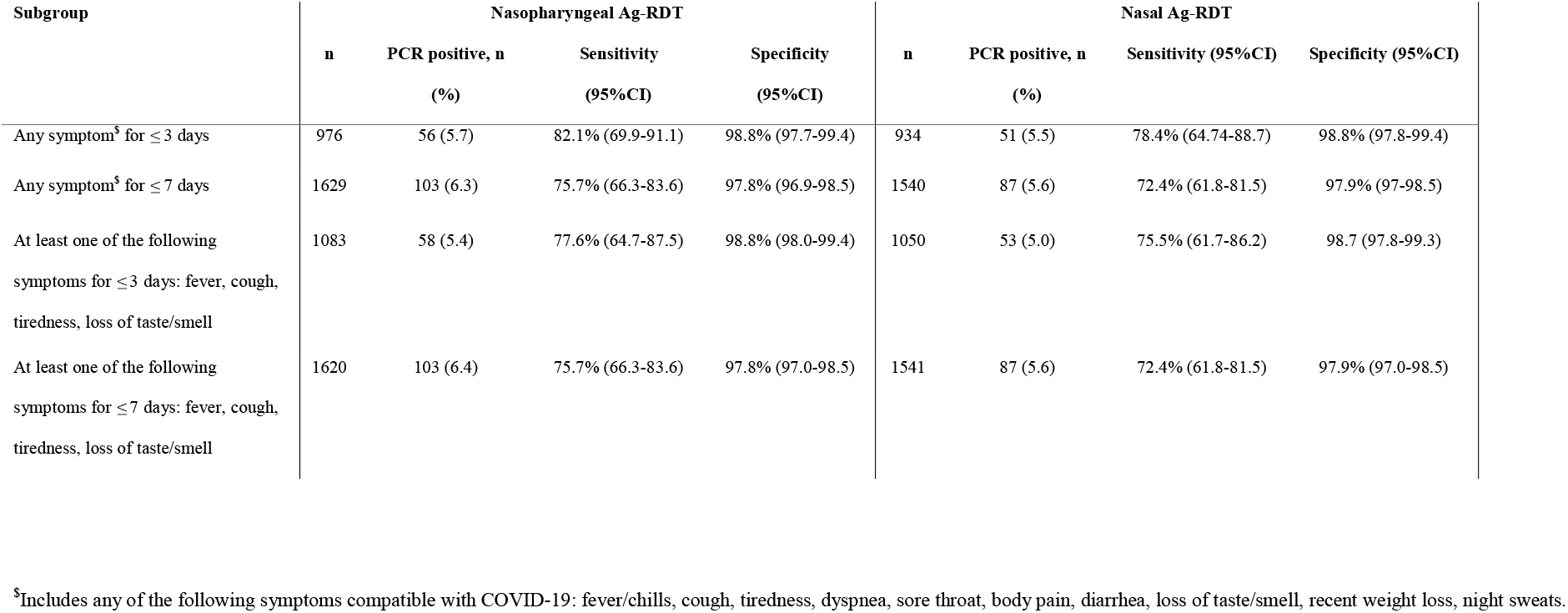
Sensitivity and Specificity of Ag-RDT on nasopharyngeal and nasal samples as compared to nasopharyngeal PCR in different subgroups

| Subgroup | Nasopharyngeal Ag-RDT |  |  |  | Nasal Ag-RDT |  |  |  |
| --- | --- | --- | --- | --- | --- | --- | --- | --- |
|  | n | PCR positive, n<br>(%) | Sensitivity<br>(95%CI) | Specificity<br>(95%CI) | n | PCR positive, n<br>(%) | Sensitivity (95%CI) | Specificity (95%CI) |
| Any symptom<sup>s</sup> for $\leq 3$ days | 976 | 56 (5.7) | 82.1% (69.9-91.1) | 98.8% (97.7-99.4) | 934 | 51 (5.5) | 78.4% (64.74-88.7) | 98.8% (97.8-99.4) |
| Any symptom<sup>s</sup> for $\leq 7$ days | 1629 | 103 (6.3) | 75.7% (66.3-83.6) | 97.8% (96.9-98.5) | 1540 | 87 (5.6) | 72.4% (61.8-81.5) | 97.9% (97-98.5) |
| At least one of the following<br>symptoms for $\leq 3$ days: fever, cough,<br>tiredness, loss of taste/smell | 1083 | 58 (5.4) | 77.6% (64.7-87.5) | 98.8% (98.0-99.4) | 1050 | 53 (5.0) | 75.5% (61.7-86.2) | 98.7 (97.8-99.3) |
| At least one of the following<br>symptoms for $\leq 7$ days: fever, cough,<br>tiredness, loss of taste/smell | 1620 | 103 (6.4) | 75.7% (66.3-83.6) | 97.8% (97.0-98.5) | 1541 | 87 (5.6) | 72.4% (61.8-81.5) | 97.9% (97.0-98.5) |

<sup>s</sup>Includes any of the following symptoms compatible with COVID-19: fever/chills, cough, tiredness, dyspnea, sore throat, body pain, diarrhea, loss of taste/smell, recent weight loss, night sweats.

### Tuberculosis diagnosis

Based on the clinical assessment, 444 of the 2131 were referred for tuberculosis work-up including Xpert Ultra testing, 24 (5.4%) had a positive Xpert Ultra sputum result and two had a “trace” result. One of the participants with a positive Xpert Ultra had a positive SARS-CoV-2 PCR at the same time point.

## Discussion

In this cross-sectional diagnostic accuracy study conducted at two rural district hospitals in Lesotho, we assessed sensitivity and specificity of the SD Biosensor STANDARD Q COVID-19 Ag-RDT in nasopharyngeal and nasal samples among persons who had either symptoms compatible with COVID-19, or reported history of contact with a person with SARS-CoV-2 during the previous 14 days. The specificity of STANDARD Q Ag-RDT in nasopharyngeal and nasal samples was above 97% as required by WHO. The sensitivity was 70% and 67% for nasopharyngeal and nasal sampling, respectively. Combining the results of the two Ag-RDT results increased sensitivity to 74%. These diagnostic performance indicators are below the WHO recommended minimum sensitivity of 80%(4), and also below the pooled sensitivity of 74.9% from 37 studies that were mainly conducted in Europe (5). Further, our study found a high agreement between nasal and nasopharyngeal sampling for Ag-RDT, indicating that for routine screening and testing in non-severely ill or asymptomatic individuals nasal sampling may replace the inconvenient nasopharyngeal swabbing. An additional, not surprising but important finding of our study is that 24 (1.1%) out of 2131 participants had pulmonary tuberculosis confirmed by a positive Xpert Ultra test on sputum (trace calls were excluded), supporting the idea of bidirectional TB/COVID-19 screening and diagnosis.

There are few studies assessing field performance for the STANDARD Q Ag-RDT on nasopharyngeal swabs in African settings, most have a small sample-size (16) (17) (18) (19) (20). Reported sensitivity ranges from 64% in a prospective study on fresh samples in Ghana (20) to 88% in a study on frozen samples in Namibia (19). Reported specificity in African studies was usually above 90% with exception from an Egypt study that reports 64.2% (17) and the above-mentioned Namibian study with 81%. In our real-world study, we could confirm findings from other groups that the sensitivity of the Ag-RDT depends on the Ct value, which is a proxy for viral load(5,21). A higher sensitivity for both RDTs was also shown in patients with ≤ 3 days since onset of any symptom compared those with to ≤ 7 days. Such improved diagnostic performance in this early stages of infection has also been noted in previous studies (5) and may be explained by high viral load in this early period after the onset of symptoms (22).

In SARS-CoV-2 testing, the nasopharyngeal swab is seen as the gold standard for sample collection. In a meta-analysis, pooled sensitivity and specificity of nasal versus nasopharyngeal PCR from eight studies was 86% (77% - 93%) and 99% (96% - 100%), respectively(9). There are few studies comparing nasal against nasopharyngeal Ag-RDT. One study using STANDARD Q Ag-RDT in 180 participants with 41 testing positive on nasopharyngeal PCR, positive percent agreement between the two sampling approaches for Ag-RDT was 93.5%(10). Similarly, another study using STANDARD Q Ag-RDT comparing anterior nasal and mid-turbinate found a positive percent agreement of 100%(23). Further head-to-head comparisons between nasal and nasopharyngeal sampling but using different Ag-RDTs came to similar results(11).

To our knowledge, our study is the largest of its kind in sub-Saharan Africa and the first providing a head-to-head comparison of nasal and nasopharyngeal Ag-RDT in such a setting. Further strengths are the prospective standardized sampling methods in a representative cohort including children in Lesotho and blinding of Ag-RDT and PCR readers to the PCR and the Ag-RDT result, respectively.

Limitations are that the study was conducted only at two clinics and that nasal and nasopharyngeal Ag-RDT were read by the same reader by visual inspection without the use of a reading device, which might have allowed for a more standardized test interpretation. Further, the previous nasal swab for Ag-RDT may have influenced the yield of the subsequent two nasopharyngeal swabs in participants with low viral secretions(24). For ethical reasons, we had to exclude critically ill patients, which may have led to a certain spectrum bias by underrepresenting patients with very advanced disease.

In conclusion, this prospective study including 2131 not critically ill participants with COVID-19 compatible symptoms or exposure to SARS-CoV-2 in Lesotho showed a rather low overall sensitivity of the STANDARD Q Ag-RDT on nasopharyngeal and nasal sampling of about 70% and 67% whereas the specificity was above 97%. Agreement between nasal and nasopharyngeal sampling for the Ag-RDT was high, suggesting that the more convenient nasal sampling may be sufficient for routine screening and testing in outpatient settings.

## Supporting information

Ethics Approval Switzerland

Ethics Approval Lesotho

## Data Availability

All data produced in the present work are contained in the manuscript

## Acknowledgements

The authors would like to acknowledge all the study participants, the clinical and laboratory staff and site investigator teams at St Charles Mission Hospital Seboche, Mokhotlong Government District Hospital and Butha-Buthe Government District Hospital. Further we would like to thank the team at the National Reference Laboratory of Lesotho for PCR analyses and the SolidarMed Lesotho team for all the logistic support.

