## Supplementary material for "Head-to-head comparison of nasal and nasopharyngeal sampling using SARS-CoV-2 rapid antigen testing in Lesotho": Ethics Approval Switzerland

EKNZ

Ethikkommission
Nordwest- und
Zentralschweiz

Präsident
Prof. Christoph Beglinger
Vizepräsidenten
Dr. Angela Frotzler
Dr. Marco Schärer

MD, MScIH, PhD Klaus Reither
Swiss Tropical and Public Health Institute
Clinical Research Unit
Department of Medicine
Socinstrasse 57
4051 Basel

Basel, 27. August 2020 / SK

**Statement of the Ethics Committee Northwest and Central Switzerland (EKNZ) according to HRA Art.51**

| | |
|---|---|
| **Statement ID** | AO_2020-00018 |
| **Statement on** | ☐ a data registry<br>☐ a biobank<br>☒ a research project outside of the scope of the HRA<br>☐ a general consent<br>☐ an ethical question |
| **Name of the data register**<br>**Name of the biobank**<br>**Title of the research project** | Mitigation strategies for communities with COVID-19 transmission in Lesotho using artificial intelligence on chest x-rays and novel rapid diagnostic tests (MistraL) |
| **Responsible Person at Main site** | MD, MScIH, PhD Klaus Reither |
| **Sponsor** | Swiss Tropical and Public Health Institute |
| **Ethics Committee** | **Headquarters /Institution** |
| **Ethics Committee Northwest and Central Switzerland EKNZ** | Swiss Tropical and Public Health Institute<br>Clinical Research Unit<br>Department of Medicine<br>Socinstrasse 57<br>4051 Basel |
| **Involved Ethikkommissionen** | None |

**Statement**

☒ positive statement

☐ statement with conditions

☐ a statement can't be given

The EKNZ has reviewed the submitted documents and can confirm that the research project fulfils the general ethical and scientific standards for research with humans (see Art. 51 Abs. 2 HRA).

The EKNZ has reviewed the project on 19. August 2020 according to the ICH GCP guideline and confirms, that the project meets all requirements for a Swiss research projects, namely:

- Scientific validity of the research question, appropriateness of the scientific design and conformity to the GCP guideline;
- Favourable benefit-risk ratio;
- Process to obtain informed consent, including the appropriateness of the period for reflection;
- Professional qualification of the research scientist involved in the project in Switzerland;
- Data protection and procedures to maintain confidentiality of the data and the samples.

Whether the project can be accepted from an ethical point of view, depends on local conditions and is the responsibility of the responsible local ethics committee. Following points could not be assessed by the EKNZ:

- Procedure and documentation for recruitment of study subjects, especially the information sheets and consent forms in the local language;
- The adequacy of the local infrastructure with regards to the best possible protection of the study subjects (material, premises, personnel etc.);
- Professional qualitifcation of the research personnel in the local country;
- Local laws and regulations.

**Fees**

**Amount:** **CHF 500.--** **Tariff code:** **6.0**

In accordance with the current swissethisc fee schedule.

**Copie to**

☐ Applicant

☒ Responsible Person at the main site MD, MScIH, PhD Klaus Reither

☐ Sponsor

☐ Representation of the sponsor in Switzerland

☐ Involved Ethikkommissionen

☐ Other

With the Committee's best wishes for the success of this project.

Yours sincerely,

i.V.

Dr. rer. biol. hum. Angela Frotzler
Vizepräsidentin der Ethikkommission
Nordwest- und Zentralschweiz / EKNZ

Attachment 1. List of documents, submitted on 29. July 2020 and 30. July 2020

| Dokument | Dok.Datum | Version |
|---|---|---|
| **1. Cover Letter** | | |
| 1-mistral-cover-letter-to-eknz-reither-burri-29-07-2020.pdf | 29/07/2020 | |
| 1bis-mistral-enclosure-list-eknz-submission-29-07-2020.pdf | 29/07/2020 | |
| **9. Participant information sheet and informed consent (ICF)** | | |
| 3-mistral-assent-children-v1-0-29-07-2020.pdf | 29/07/2020 | 1.0 |
| 3-mistral-icf-children-v1-0-29-07-2020.pdf | 29/07/2020 | 1.0 |
| 3-mistral-icf-adults-v1-0-29-07-2020.pdf | 29/07/2020 | 1.0 |
| **10. Study plan (protocol)** | | |
| 4-mistral-study-protocol-v1-0-29-07-2020.pdf | 29/07/2020 | 1.0 |
| **11. Investigator's CV** | | |
| 6-mistral-cvs.7z | 29/07/2020 | NA |
| **12. Agreement between sponsor/commissioned institution/grant provider** | | |
| 9-mistral-funding-agreement-ftc-12-05-2020.pdf | 12/05/2020 | NA |
| **20. Varia** | | |
| 5-mistral-crfs-draft-v1-0-29-7-2020.pdf | 29/07/2020 | 1.0 |
