## Supplementary material for "Head-to-head comparison of nasal and nasopharyngeal sampling using SARS-CoV-2 rapid antigen testing in Lesotho": Ethics Approval Lesotho

Ministry of Health
P.O. Box 514
Maseru 100

**Category of Review:**
[x] Initial Review
[ ] Continuing Annual Review
[ ] Amendment/Modification
[ ] Reactivation
[ ] Serious Adverse Event
[ ] Other ________________

**REF: ID 107-2020**
Date: November 13, 2020
To
**Dr Josephine Muhairwe,**
Lesotho Site Investigator
Country Director,
SolidarMed Lesotho

Dear **Dr Josephine**

**RE: "Mitigation strategies for communities with COVID-19 transmission in Lesotho using artificial intelligence on chest x-rays and novel rapid diagnostic tests." (MistraL).**

This is to inform you that the Ministry of Health Research and Ethics Committee reviewed and **APPROVED** the above named protocol and hereby authorizes you to conduct the study according to the activities and population specified in the protocol. Departure from the approved protocol will constitute a breach of this permission.

This approval includes review of the following attachments:
[x] MistraL, Study protocol, v1.1 19.10.2020
[x] **Informed consent form**: Mistral ICF adult's English v1.1 19.10.2020, Mistral ICF adults Sesotho v1.1 19.10.2020, Mistral ICF Children English v1.1 19.10.2020, Mistral ICF Children Sesotho v1.1 19.10.2020, Mistral Assent Children English v1.1 19.10.2020, and Mistral Assent Children Sesotho v1.1 19.10.2020
[x] **Participant materials**: MistraL, CRFs, draft v1.0, 29.7.2020
**[x] Other materials**: Letter of request dated 1st September 2020, Letter of Response to NH-REC queries on MistraL project ID 107-2020 dated 19th October 2020, Statement of the Ethics Committee Northwest and central Switzerland (EKNZ) dated 12 August 2020 Investigator's **CVs, Investigators**(Josephine Muhairwe,Keelin Murphy, Klaus Reither, Lucia Gonzalez, Morten Ruhwald, Nklaus Labhardt, Samuel Schumacher,Tracy Glass, B Van Ginneken), **GCP certificates**(Josephine Muhairwe,Keelin Murphy, Klaus Reither, Lucia Gonzalez, Morten Ruhwald, Nklaus Labhardt, Samuel Schumacher,Tracy Glass, B Van Ginneken),

This approval is **VALID** until November 16, 2021.

Please note that an annual report and request for renewal, if applicable, must be submitted at least 6 weeks before the expiry date.

All serious adverse events associated with this study must be reported promptly to the MOH Research and Ethics Committee. Any modifications to the approved protocol or consent forms must be submitted to the committee prior to implementation of any changes.

We look forward to receiving your progress reports and a final report at the end of the study. If you have any questions, please contact the Research and Ethics Committee at (or) 59037919/58800246.

Sincerely,

**Dr. 'Malitaba Litaba (a.i)**
Director General Health Services

**Dr. Jill Sanders**
Co- Chairperson National Health Research Ethics Committee (NH-REC)
